# Care delivery in the context of district mental healthcare plans (DMHP) in Ghana: experiences of primary health care workers and service users

**DOI:** 10.1101/2024.06.30.24309723

**Authors:** L Sakyi, KA Ae-Ngibise, L Adwan-Kamara, Ben Weobong, Crick Lund

**Author notes:** **Corresponding author** Lionel Sakyi /.

## Abstract

**Background:** The integration of mental healthcare into primary healthcare services is an effective way to addressing the substantial treatment gap for mental health conditions in low- and middle-income (LMIC) countries. In Ghana, District Mental Healthcare Plans (DMHPs) were developed and implemented in three districts over a 2-year period. This study aimed to explore the perceptions and experiences of mental health service users and primary healthcare workers regarding the implementation of the DMHPs.

**Methods:** The study employed a qualitative design involving in-depth interviews with 32 service users and 28 primary healthcare providers in the three demonstration districts. Participants were purposively sampled. Interview data were analysed using reflexive thematic analysis combining inductive and deductive approaches.

**Results:** Three main themes were identified: 1) Factors supporting DMHP implementation, including capacity building, collaboration, awareness creation, and acceptability; 2) Challenges impacting DMHP implementation, such as inadequate resources and medication shortages; and 3) Impacts of the DMHPs, including improved access to care, reduced relapse, enhanced daily functioning, and reduced stigma. Some district-level variations were noted in the intensity of challenges and impacts.

**Conclusion:** The DMHPs showed promise in improving mental healthcare in primary care settings in Ghana. However, addressing resource constraints and medication shortages, and sustaining capacity building and awareness creation efforts, will be crucial for successful scale-up. The perspectives of service users and healthcare providers offer valuable insights for policy makers and practitioners aiming to enhance integrated mental healthcare.

**Strengths and limitations of this study:**

⇒ This study is the first in Ghana to explore in-depth the experiences of healthcare workers and service users in implementation of DMHPs.
⇒ Using qualitative design provide more nuanced understanding of factors supporting implementation of DMHP which would have been more difficult to do using quantitative methods.
⇒ The study reported from a relatively small sample size from only 3 districts from 261 districts in Ghana. While this sample size provided valuable insights, it may not cover the wide and diverse experiences of healthcare workers and service users involved in the DMHPs.

## INTRODUCTION

Mental health conditions are a major public health burden globally and account for 16% of disability-adjusted life years worldwide.^1^ The situation has been exacerbated by COVID-19 pandemic with an increase in mental, neurological and substance use disorders such as depression and anxiety.^2^ Despite an estimated 80% of people with mental health conditions living in low- and middle-income countries (LMIC), access to mental health services is limited due to socioeconomic disparities, inadequate human and financial resources, cultural stigmatisation, lack of policy priority and insufficient mental health infrastructure.^3^ In Ghana, the trend is similar and a mental healthcare gap of between 94-98% has been reported.^4,5^ Inability of people with mental health conditions to access mental health services may exacerbate their conditions and impair their daily functioning.^6^

The integration of mental health services in primary healthcare services has proven to be one of the most effective ways of improving access to mental healthcare services.^7,8^ The benefits of integrating mental health services into primary healthcare include reducing the care gap and the impact of mental health conditions on individuals.^9,10^

Ghana Somubi Dwumadie (Ghana Participation Programme) is a four-year disability programme in Ghana, with a specific focus on mental health. The programme seeks to provide support and generate evidence to inform the effectiveness of mental health programmes and interventions in Ghana (https://www.ghanasomubi.com/).

As part of the strategy to improve mental healthcare access in Ghana, Ghana Somubi Dwumadie has worked with local stakeholders to develop and implement district mental healthcare plans (DMHP) in three demonstration districts in Ghana. Using the Programme for Improving Mental HealthcarE (PRIME) approach^7^, the DMHPs were designed to raise mental health awareness, improve detection, treatment, and recovery, as well as improve overall functioning of service users. One of the components of the DMHPs was implementing interventions that would enhance the integration of mental health services at primary care level. This included the training of healthcare workers and community health volunteers on the WHO mhGAP intervention guide to detect and manage MNS conditions as well as provide psychosocial support to service users.^11^ The implementation steps as well as the evaluation have been explained in an earlier publication.^12^ Despite the emerging evidence on the process of developing and implementing DMHPs, there is a gap in our knowledge regarding the perceptions and experiences of mental health service users and primary healthcare workers involved in this process. Understanding the perspectives and experiences of primary healthcare providers and service users in the implementation of the mental healthcare plan is important for identifying acceptability, feasibility, areas of success, challenges, and opportunities for improvement in mental healthcare delivery. The perspectives of service users are vital if we are to integrate mental healthcare into primary care in Ghana.^13^

The aim of this paper is to explore the perceptions and experiences of mental health service users and healthcare workers regarding the implementation of DMHPs in three district demonstration sites in Ghana. Specifically, we set out to examine the factors supporting the DMHP implementation; explore challenges impacting successful implementation of the DMHP; and examine the impact of DMHP in the three districts.

## METHODS AND DESIGN

### Study design

The study employed a qualitative research design using thematic analysis to understand the perceptions and experiences of both mental health service users and primary healthcare workers regarding the implementation of the DMHPs.^14^ Qualitative thematic analysis was considered appropriate as it facilitates a rigorous in-depth exploration of themes in qualitative data, in this case regarding participants’ perception of the DMHPs. The study used both inductive and deductive thematic content analysis approaches.^14^

### Settings

The study was conducted in three demonstration districts where the DMHPs were being implemented as part of efforts to improve access to mental healthcare services in the country. These districts are Anloga in the Volta Region, Asunafo North in the Ahafo Region, and Bongo in the Upper East Region. The districts were selected through an extensive consultation process with local and national stakeholders in which a framework for DMHP was outlined. The rationale and criteria for the selection of these three districts as demonstration sites have been described in other publications.^5,12^ A situational analysis was conducted to assess the healthcare system and available resources in the districts and the findings have been reported previously ^5^. After selecting the districts, five health facilities in each district were randomly selected from a group of 6-10 facilities with high outpatient department (OPD) attendance and the presence of physician assistants, mental health nurses, or midwives. On average, these facilities had at least 48 OPD visits per day. The 15 health facilities included 10 health centres, 2 hospitals, 2 Community Health Planning and Services and 1 clinic.^4^

### Participants

In-depth interviews were conducted with primary healthcare workers who played key roles in the delivery of care in the demonstration districts. The composition of the healthcare workers included midwives, general nurses, physician assistants, enrolled nurses and mental health specialists. In-depth interviews were conducted with service users who are 18 years and above and have been receiving healthcare for the past year at the 15 health facilities (5 in each district) that were selected during the implementation of the mental healthcare plans in the three demonstration districts. The facility selection process and methodology has been extensively covered in another paper ^4^.

### Sampling and recruitment procedure

Purposive sampling was used to recruit all study participants. Participant recruitment was facilitated by two of the researchers (LS and KA) and the mental health coordinators in the districts. Before the start of the recruitment, meetings were held with the district directors from the three districts to brief them on the purpose of the research. With the help of the district mental health coordinators, healthcare workers who had been part of the implementation of the DMHPs were identified. They were then contacted by their respective district mental health coordinators to inform them of the researchers’ intention to interview them. Contacts with identified potential participants were made by the researchers through telephone calls and interview dates and times were scheduled.

With regards to service users, the district mental health coordinators contacted the department heads of selected healthcare facilities to identify service users who had received treatment in the past year since the implementation of the DMHPs in the districts. The identification was done through the review of folders of service users to ascertain period of treatment. The selected potential participants were then contacted by the district mental health coordinators to inform them of the researchers’ plan to interview them for the study. Contacts were made either directly to service users or their caregivers by the research team. Following this, dates and times were arranged, and interviews were conducted by researcher assistants and two of the researchers with the assistance of the district mental health coordinators. The district mental health coordinators assisted in identifying participants and at no point did they interview participants. All study participants provided voluntary informed consent to participate. Recruitment of both healthcare workers and service users was done until thematic saturation was reached.

### Data collection

In-depth interviews were chosen as the primary method of data collection to facilitate deeper understanding of participants’ experience of care delivery in the context of DMHPs. Interviews were conducted in two phases. In Phase 1 of interviews, 3 research assistants were trained and conducted interviews with both service users and healthcare workers. Phase 2 of the interviews were conducted face to face by LS and KA across the three districts. The Phase 1 interviews allowed for the identification of main themes, which were explored in more detail by LS and KA in phase 2. The use of RAs was important because they were less involved in the design and evaluation of the DMHPs, and therefore provided a greater degree of objectivity. Interviews were conducted in both English and the local dialects (Ewe, Frafra and Twi) by the researchers. Data collection was conducted in places such as health facilities as well as homes of service users. The interview guide was developed by the research team guided by the research questions of the study with probes and follow up questions to seek clarifications (See online supplementary file). Key thematic areas that were explored in the interviews for service users included acceptability of healthcare provided, experience of stigma and discrimination, and barriers accessing and adhering to treatment. For healthcare workers, thematic areas explored included lessons learned from implementing mental healthcare plans, acceptability of delivering mental healthcare plans, availability of resources for implementation of mental healthcare plan, social protection, economic empowerment and training and supervision. The interview sessions were audio recorded. The interviews lasted between 15 and 20 min on average.

### Data analysis

The audio data from the in-depth interviews were extracted from the recorder and organised into two main folders with each representing healthcare workers and service users. To ensure the anonymity of participants, the file names of both the healthcare workers and service users’ interviews were replaced with different identifiers. All the interviews were transcribed verbatim into English by skilled transcribers with expertise in the three languages (Twi, Frafra and Ewe) in which the interviews were conducted. The Twi and Frafra transcripts were double checked by two researchers (LS and KA). The Ewe transcripts were double checked by the district mental health coordinator.

A reflexive thematic analysis^14^ using inductive and deductive approaches was used to gain a better understanding of healthcare providers’ and service users’ perspectives on critical factors influencing care delivery in the districts. The deductive approach was used to create the main themes and sub-themes from the research questions in the semi-structured interview guide. The inductive approach ensured that all the themes that emerged from the transcripts that did not exist in the deductive codes were captured. Analysis proceeded through six stages: (1) reading the transcripts closely to gain familiarity with the text; (2) generating initial codes to capture the range of views on the experiences of healthcare workers and service users; (3) collating the codes into themes and gathering all the text related to the themes and subthemes; (4) reviewing the themes and subthemes and refining them; (5) defining and naming the themes that have been identified; and (6) selection of vivid examples that capture the essence of the themes and subthemes to write the analysis report.

### Reflexivity Statement

As researchers involved in the implementation of the DMHPs being studied, it is important that we reflect on our roles and potential biases. Our close involvement in designing and implementing the DMHPs may have influenced our perceptions and interpretations of participants’ experiences. To mitigate this, we employed strategies such as having the phase 1 of the interviews conducted by researcher assistants who were less directly involved in DMHP implementation. We also made efforts to probe for both positive and negative experiences during the interviews.

However, participants’ responses may still have been affected by social desirability bias, as they might have wanted to provide positive feedback to researchers seen as connected to the DMHP implementation. Our backgrounds as mental health researchers and practitioners also inevitably shape our perspectives. We may hold assumptions about the importance and appropriateness of integrating mental healthcare into primary care based on our disciplinary training and previous research.

To ensure credibility, we aimed to engage in reflexivity throughout the research process - critically examining our own positioning, biases and assumptions. Triangulating the interview data with health facility and DMHP records could have provided further validation but was not feasible within the scope of this study. Including more independent researchers in the data analysis could have helped counterbalance potential biases among those of us involved in DMHP implementation.

Despite these limitations, we believe this study provides important insights into the experiences of service users and healthcare workers during the early implementation of the DMHPs.

## RESULTS

### Characteristics of the study participants

A total of 60 participants (32 service users and 28 healthcare workers) were recruited and interviewed about their perspectives and experiences in the implementation of the DMHPs in the three demonstration districts. Table 1 provides further details about the characteristics of participants.

**Table 1:** Demographic characteristics of participants.

| Characteristics | Bongo | Asunafo<br>North | Anloga | Total |
| --- | --- | --- | --- | --- |
| Service users | 11 | 10 | 11 | 32 |
| Male | 7 | 7 | 5 | 19 |
| Female | 4 | 3 | 6 | 13 |
| **Healthcare workers** | 8 | 10 | 10 | 28 |
| Male | 4 | 4 | 4 | 12 |
| Female | 4 | 6 | 6 | 16 |
| Subtotal | 19 | 20 | 21 | 60 |
| **Designation** |  |  |  |  |
| Community Mental Health Officer | 3 | 2 | 2 | 7 |
| Midwife | 2 | 2 | 3 | 7 |
| Registered Mental Health Nurse | 2 | - | - | 2 |
| Physician Assistant | 1 | - | - | 1 |
| Registered Community Nurse | - | 3 | 2 | 5 |
| Registered General Nurse | - | 1 | - | 1 |
| Enrolled nurse | - | 2 | 2 | 4 |
| Senior Nurse Officer | - | - | 1 | 1 |
| Total | 19 | 20 | 21 | 60 |

### Summary of key themes

The study sought to explore the perception and experiences of healthcare workers and service users in the implementation of the mental healthcare plans in the three demonstration districts. Three main themes emerged from the analysis of the data. These themes were factors supporting district mental healthcare implementation, challenges impacting successful implementation of DMHPs and the impact of the plans. Table 2 describes each of the themes and the subthemes.

**Table 2:**
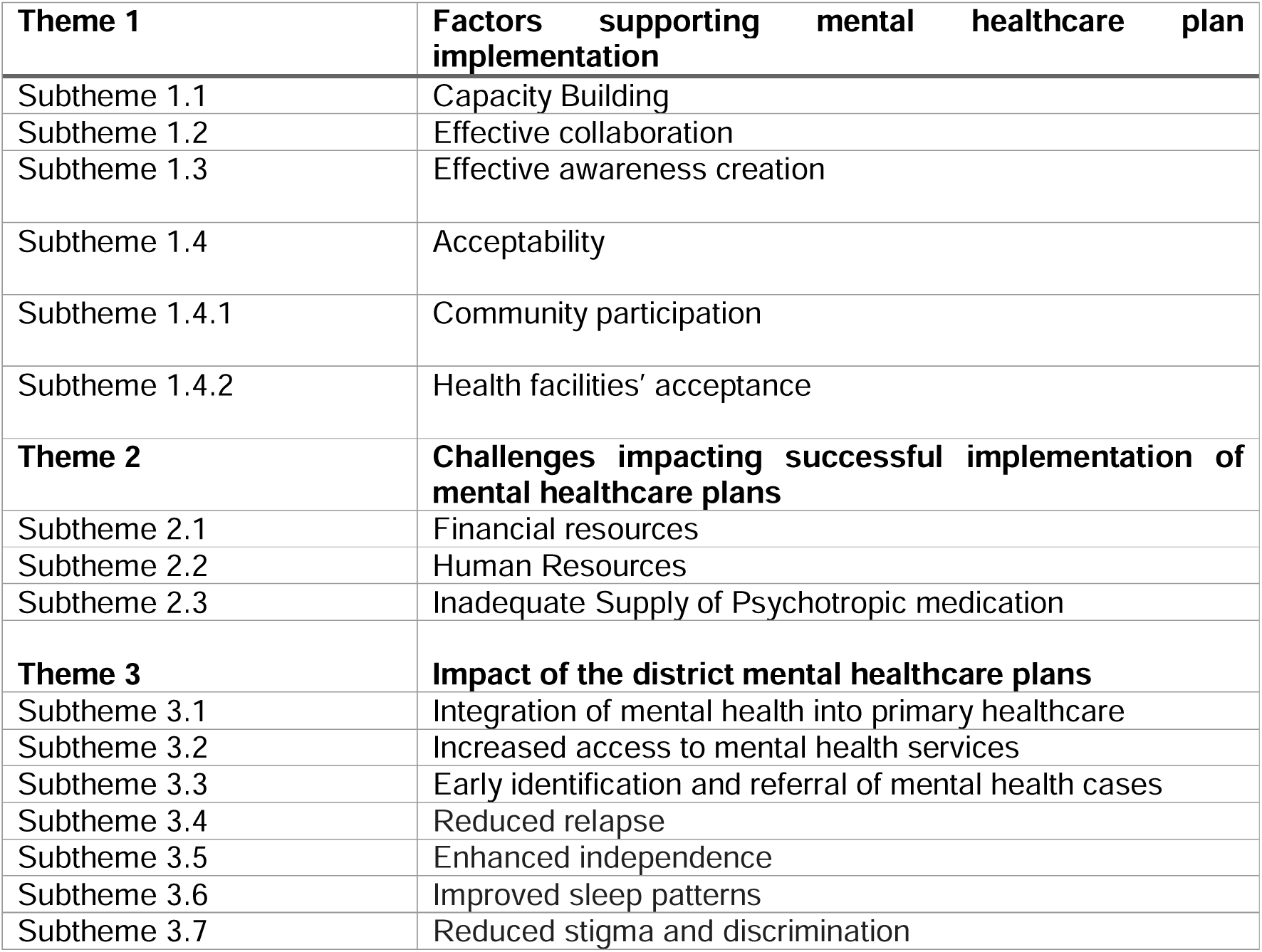
Healthcare workers’ and service users’ experiences of district mental healthcare plan implementation.

| **Theme 1** | **Factors supporting mental healthcare plan implementation** |
| --- | --- |
| Subtheme 1.1 | Capacity Building |
| Subtheme 1.2 | Effective collaboration |
| Subtheme 1.3 | Effective awareness creation |
| Subtheme 1.4 | Acceptability |
| Subtheme 1.4.1 | Community participation |
| Subtheme 1.4.2 | Health facilities' acceptance |
| **Theme 2** | **Challenges impacting successful implementation of mental healthcare plans** |
| Subtheme 2.1 | Financial resources |
| Subtheme 2.2 | Human Resources |
| Subtheme 2.3 | Inadequate Supply of Psychotropic medication |
| **Theme 3** | **Impact of the district mental healthcare plans** |
| Subtheme 3.1 | Integration of mental health into primary healthcare |
| Subtheme 3.2 | Increased access to mental health services |
| Subtheme 3.3 | Early identification and referral of mental health cases |
| Subtheme 3.4 | Reduced relapse |
| Subtheme 3.5 | Enhanced independence |
| Subtheme 3.6 | Improved sleep patterns |
| Subtheme 3.7 | Reduced stigma and discrimination |

#### Theme 1: Factors supporting district mental healthcare plan implementation

Factors that were identified as supporting the implementation of the DMHPs in the districts were capacity building, effective collaboration, effective awareness creation and acceptability of the district mental healthcare plan. The theme of acceptability was categorised into two subthemes: community participation and health facilities’ acceptance.

#### Subtheme 1.1: Capacity Building

One significant factor that was reported to positively impact district mental healthcare delivery was the capacity building of healthcare workers within the districts, as part of the implementation of the DMHPs. Respondents discussed the importance of targeted training and the role it had played in empowering healthcare workers and community volunteers in providing effective mental healthcare delivery to service users. The capacity building not only improved the skills and knowledge in mental health detection of healthcare workers but also increased their awareness and prioritization of mental healthcare. Respondents specifically mentioned that the training has equipped them with skills to identify and manage mental health cases.

> “The training you provided on mental health to us at the sub-district have really equipped and helped us on cases involving mental health and even […] how to identify and treat through this programme.” Registered General Nurse ID 015
>
> “Even though before your coming, it was part of the service we render, your presence has also empowered us in a way to pay special attention to mental health in the sense that you started the team drawing the plan for the district, proceeded by training staff on the mhGAP and then you came back to train our volunteers who will support work at the community level.” Senior Nursing officer, ID 010

Respondents discussed the importance of training and empowering community health volunteers as part of the capacity building efforts in implementing the DMHPs. They explained that community health volunteers had played an important role in providing education to community members after receiving training in mental health detection and referral.

> “They are happy. One of the major community goals is that we work with the community health volunteer. Majority of them too were brought on board and we trained them on case identification and then referral. So once they went back home they continued the education process and have started referring cases within the community to us…” Community Mental Health Officer, ID 009

#### Subtheme 1.2: Effective collaboration

Effective collaboration between different stakeholders in improving mental health service delivery and support for individuals with mental health conditions was a common feature of the DMHP implementation in all the districts. For example, the district health management team collaborated with the assembly and the social welfare department to distribute monetary support to individuals in need. This financial assistance can play a role in supporting the well-being and recovery of individuals with mental health conditions, particularly those facing socioeconomic challenges.

> “At first when you go to other members on the operational team who are not health related, once you tell them the problem, they usually say it is a health-related case. But now since they have been part of the implementation process throughout the beginning, they understand certain challenges facing the health sector and they are willing to come to our aid especially with the social welfare and the support services. That has been very key in the implementation process.” Registered Mental Health Nurse, ID 020

#### Subtheme 1.3: Effective awareness creation

Awareness creation was identified as one of the important factors in the successful implementation of the DMHPs. Bongo district demonstrated a more intensive outreach and awareness efforts compared to the rest of the districts. Respondents emphasised the effectiveness of outreach services, such as joining the Reproductive and Child Health (RCH) activities, conducting health education at Outpatient Departments (OPD), and organising community durbars (engagement meetings). These activities provided them with the opportunities to directly engage with community members, educate them about mental health, and promote the availability of services at health facilities. The healthcare workers also observed how increased awareness during the implementation of the mental healthcare plans had led to improved utilisation of mental health services. The awareness creation was achieved through collaborations with community leaders and opinion leaders, through media and public education.

> “What actually works for us. Mostly it’s on our outreach services. We go with this…our RCH when they are going for weighing. We get the mothers, we talk to them, and then I think that has been very good way of helping us improve mental healthcare delivery. And then we normally do health educations at the OPD every morning and it’s also working for us. It is at least contributing to people understanding mental health as a whole so that they can just accept it and then embrace it.” Enrolled Nurse, ID 005

#### Subtheme 1.4: Acceptability of the district mental healthcare plan

As key stakeholders and implementers of the DMHPs, healthcare workers provided their perceptions of the extent to which the mental healthcare plans were considered satisfactory or suitable. The subthemes generated were categorised under community participation and health facilities acceptance.

#### Subtheme 1.4.1: Community participation

Healthcare workers interviewed by the study explained that community involvement and engagement had been key components in the implementation of the mental healthcare plans in the districts. They specifically highlighted the role played by the Assembly person, who are local government representatives, in taking leads in organizing people to attend durbars organised as part of the implementation of the DMHP. Evidence of communities’ acceptance of the mental healthcare plan is the fact that healthcare workers are given free airtime to discuss mental health issues on the radio and raise awareness, a critical aspect of the plan’s objective of reducing mental health stigma and discrimination.

> “Yes, yes, yes they have accepted it. I will congratulate the Assembly Men. They are doing marvellously well. Sometimes you try to organize a durbar and they will take it up, organize people to come and listen to the talk and the advice you are giving them. So the community as a whole has also supported us” Community Mental Health Officer, ID 07

#### Subtheme 1.4.2: Health facilities’ acceptance

Interviews with some of the healthcare workers showed that many of them actively participated in the planning and implementation of the DMHPs. Some who are also members of the operation team took ownership of their roles and responsibilities within the plans. Healthcare workers acknowledged that the introduction of the mental healthcare plan has been a valuable tool that had helped improve mental health services in the districts.

> “Yes, the plan has been accepted that is why we are making significant progress when it comes to mental health. When you review a few selected folders in some facilities, you will realize that some of the questions we ask clients when they come, which we document in the folder, also give us that signal that we are actually screening for probable cases.” Midwife, ID 03
>
> “yes, for the healthcare providers, we have always been looking for means to deliver the best of services to the clients, so with the district mental healthcare plans, you could clearly see that it is a work plan, like a road map for us to ride on in order to be able to achieve whatever we aim at, not just for Somubi, but for Ghana health service too.” Community Mental Health Officer, ID 12

#### Theme 2: Challenges impacting successful implementation of mental healthcare plans

Under this theme, three sub-themes were identified from the data. The challenges were thematised into financial resources, human resources, and inadequate supply of psychotropic medication.

#### Subtheme 2.1: Financial resources

The most significant factor influencing mental healthcare delivery according to all the healthcare workers in this study is inadequate financial resources and inadequate financial support at the national level, which hampered the overall mental healthcare delivery. More respondents in Anloga district reported inadequate financial resources as constraints compared to respondents from Bongo and Asunafo district.

> “No, that is a big challenge. Considering the financial position of the nation as a whole, releasing funds now is a huge problem. This has in turn crippled the [district mental healthcare] plan to some extent.” Senior Nursing officer, ID 010

Outreach programmes were considered to be crucial for creating awareness, identifying cases, and providing mental health services to communities, especially those in remote areas. However, the success of these programmes relied heavily on the availability of transportation and the necessary financial resources to support them. The lack of money to purchase fuel hindered the healthcare workers’ ability to effectively carry out their plans. This situation posed a challenge to the implementation of the DMHP. One healthcare provider commented:

> “Even getting money these days to buy fuel into the available vehicle is also a problem, which poses a challenge to the outreach programs.” Registered Community Nurse, ID 028

#### Subtheme 2.2: Human Resources

One of the critical challenges in mental healthcare delivery was the shortage of healthcare workers in the districts. The shortage of mental health professionals was one of the challenges faced by many districts in implementing DMHPs. This shortage was seen to put pressure on the existing mental health staff and limit the district’s capacity to provide adequate mental health services to the population.

> “Yes, the district has adequate number of health personnel, but for the mental health professionals, they are very limited. Only four of them that are in the district, taking care of a population close to 99,000. I think we need to do something about that one. For the health professionals, they are adequate, but for the mental health professionals, they are woefully inadequate.” Registered Mental Health Nurse, ID 022

#### Subtheme 2.3: Inadequate Supply of Psychotropic medication

All respondents mentioned inadequate supply of psychotropic medication as one of the main challenges impacting the implementation of the DMHPs in the districts. The lack of essential medicines was reported to impact the ability of healthcare workers to provide optimal care for individuals with mental health conditions. Health facilities from Anloga and Asunafo North districts experienced acute shortage of psychotropic medicines compared to Bongo. Most service users interviewed expressed their frustrations at the lack of medication during their visits to their respective health facilities. Most of the times they are directed by service providers to seek their medications from private pharmacies or other healthcare facilities. This situation not only delays their access to necessary treatment but also poses a financial burden if they are required to purchase the medications from private pharmacies or drug stores.

> “For the resources I think that if a staff or clinician identifies a case and the case needs to be managed with medication and then the medications are not there, that clinician is limited as to the quality of care he or she can provide for the client.” Community mental Health Officer, ID 007
>
> “I would say that some of the barriers are lack of injections and drugs they are the barriers sometimes when I get there the nurse always complained about non-availability of injections and drugs so they direct me to different health facility to check whether I would get it or not but now I think is getting better a little bit” Service user, ID 019

#### Theme 3: Impact of the district mental healthcare plans

Regarding the impact of the DMHPs, the themes mentioned by healthcare workers included integration of mental health into primary healthcare, early identification, and referral of mental health cases. From the perspectives of service users, participants mentioned improved access to mental health services, reduced relapse, enhanced independence, improved sleep patterns and reduced stigma and discrimination as impacts of the DMHPs.

#### Subtheme 3.1: Integration of mental health into primary healthcare

Healthcare workers who were interviewed in all 3 districts stated that one of the impacts the implementation of the mental healthcare plan was the integration of mental health into primary healthcare. Participants explained that mental health was not fully incorporated into the different units of the healthcare system, and the involvement of mental health in primary healthcare activities was limited. However, the implementation of the DMHPs has facilitated the integration of mental health services into the regular services provided at the primary care level.

> “To me, I came in when they had already started the project, and the seven months that I have been in the program, it’s been very good. Why am I saying this? I say this because gone were the days when things were being done in terms of building to incorporate mental health into our different units of which their involvement into primary healthcare activities was not seen. So, looking at it in that way, it has helped us to integrate these services into our normal services.” Senior Nursing officer, ID 010

#### Subtheme 3.2: Improved access to mental health services

Some service users acknowledged the increased availability and accessibility of mental health services in their communities or nearby areas due to the implementation of the DMHPs. They believe that the DMHP had brought mental health services closer to the community, made them more accessible and responsive, and introduced outreach efforts to reach individuals who may face challenges in physically visiting the healthcare facility.

> “The facility is closer to us, and it is always accessible to us. We always call on them at any time, they are also trying to come to us and give help. They even sometimes come home when I am not able to come to hospital”. **Service user, ID 005**

#### Subtheme 3.3: Early identification and referral of mental health cases

The healthcare workers indicated that instead of waiting for individuals to seek help themselves, they are now able to recognize potential mental health issues even when patients present with physical symptoms at the general health facilities. One participant provide an explanation for the change, as follows:

> “Now I will say that we are able to pick up cases without waiting for them to come because, you know there are many people with mental health conditions who even go to the general side and present some physical symptoms but once these different categories of health personnel were trained [mhGAP training] they were able to pick up this cases right from there so I will say early identification of cases is one of the positive side of it.” Community Mental Health Officer, ID 001

The healthcare workers further indicated that there had been an increase in referral cases due to the training provided to non-mental health staff and community health volunteers.

> “I think there has been many referral cases from the sub districts because of the training e.g., the non-mental health staffs, now they are able to identify cases even the community health volunteers are referring cases. So you could see that clients coming to the facility has increased as compared to earlier on.” Community Mental Health Officer, ID 025

#### Subtheme 3.4: Reduced relapse

Interviews with some service users affirmed improvements in their mental health conditions. They indicated they had noticed a reduction in relapses and overall improvements in their health conditions because of the implementation of the DMHPs. They compared their current state to their previous state and suggested an overall improvement which they attributed to the effectiveness of the treatment they had been receiving from healthcare workers:

> “Yes of course, I feel my health condition has improved because the way I behaved previously I do not behave as such so for now I would say it has improved”. Service user, ID 007.
>
> “Yes, please it has improved. At first, I have been taking the injection, and this did not work for me well, and I talked to them, and they changed it to tablet Olanzapine, and now I am able to do my daily work.” Service user, ID 15.

#### Subtheme 3.5: Enhanced Independence in daily tasks

Twelve service users who had access to treatment reported an increased ability to perform daily tasks independently. This was attributed to the treatment they were receiving during the DMHP implementation. Prior to the treatment, service users’ mental health conditions were reported to have impeded their ability to function and work independently. However, the medicines they were receiving now were perceived as contributing to their recovery and independence. This was articulated by a service user:

> “At first it was very difficult to even have a chat with me but now I am able to do my own things like cooking, wash my own clothes unlike at first where I was unable to do anything by myself.” Service user, ID 008

#### Subtheme 3.6: Improved sleep patterns

One of the impacts of service users receiving effective medication is the improvement in their sleep patterns. A number of service users reported improved sleep patterns as a sign of their improved condition. This was attributed to the mental health services they had been receiving from healthcare workers. A participant had this to say about their sleep pattern experience:

> “…is good now, at first even to sleep they would’ve to tire me down for me to be calm before I could sleep but now I can sleep myself without any force from anyone, I sleep normal now when I feel to do so.” Service user 011

#### Subtheme 6: Reduced Stigma and Discrimination

A few service users mentioned a reduction in stigma and discrimination, either in their communities or at the healthcare facilities, potentially due to increased awareness and sensitization efforts accompanying the DMHP implementation.

> “I would say that now the public perception about me has reduced because I don’t behave like how I use to do before so I can now say my health condition has improve so the public perception about me has reduced totally.” Service User 007

## DISCUSSION

The study sought the perspectives and experiences of primary healthcare providers and service users during the implementation of DMHPs in demonstration sites in Anloga, Asunafo North and Bongo districts in Ghana. Healthcare workers and service users reported a range of factors supporting district mental healthcare implementation, challenges impacting implementation of DMHPs and the impacts of the DMHPs.

While healthcare workers mentioned inadequate supply of psychotropic medicines as one major challenge, there was differences in medication availability across districts. Health facilities from Anloga and Asunafo North districts experienced more acute shortage of psychotropic medicines compared to Bongo. While Bongo also experienced shortage in psychotropic medicine, their ability to leverage on external support may have contributed to this difference compared to the Anloga and Asunafo North districts that relied on existing budgets as reported in our earlier publication.^12^ There were varied level of logistical and financial resources constraints across the districts. More respondents in Anloga district reported financial and logistical resources compared to respondents from Bongo and Asunafo district. Over the 2-year period of DMHP implementation, there was a 159% average monthly increase in mental health care service utilisation rates in general healthcare facilities in Anloga district compared to a 106% increase in Bongo district and 63% increase in Asunafo North district as reported in our other study ^12^.

Service users reported improved access to mental health services, reduced relapse rates, improved independence in daily tasks, better sleep patterns, and decreased stigma and discrimination. Healthcare workers reiterated these sentiments, noting the increased integration of mental health into primary healthcare, improved early identification and referral of mental health cases, and the positive effects of capacity-building. These outcomes suggest that the DMHPs has effectively impacted some important areas of mental healthcare delivery in the districts.

### Findings compared with previous literature

The valuable effect of capacity building of healthcare workers on facilitating integration of mental healthcare into primary care has been highlighted as one critical factor in implementation of mental healthcare plans in India^16^ corroborating the findings from the study. Effective collaboration between government agencies has proven to be one of the factors supporting the successful implementation of the DMHPs in our study. This finding is consistent with studies that used integrated mental health approach to improve access to mental health services in Kenya.^17^ Positive support from health facilities enhanced the integration of mental health into primary healthcare system and this was achieved through active involvement of healthcare workers as evidenced in other mental health integration programmes in Lebanon.^18^ This is a crucial factor in ensuring improved access to mental healthcare.

One of the emerging challenges highlighted in this study was the consistent inadequate financial and human resources support by the central government, the main stakeholder in health in Ghana. This continues to be a barrier in effective care delivery of mental health services in Ghana. Mental healthcare in LMICs has consistently been hampered by these critical issues.^18–20^ Additionally, inadequate supply of psychotropic medications was identified as a major barrier in people with mental health conditions receiving effective mental healthcare delivery. Factors include centralisation of procurement process, non-prioritisation of psychotropic medications as well as financing of psychotropic medicines.^21–24^ Evidence within the study suggest similar psychotropic medication challenges across all the districts. This challenge continues to impact mental health service delivery as service users are sometimes sent to buy medications from nearby pharmacies which put financial constraints on them. Currently, the Government of Ghana is working to is working on making outpatient treatment for some MH conditions available on National Health Insurance Scheme.^24^

Service users’ reports of overall improved condition mirrors other PRIME district mental healthcare plans, in which cohort studies showed higher response rates, early remission, and recovery among patients with depression who received care during implementation of the district plan in Ethiopia, India, Nepal, South Africa and Uganda.^3,25^ One of the objectives of the implementation of the district mental healthcare plans was to facilitate the integration of mental healthcare into the primary healthcare system in the three districts. Healthcare workers confirmed a noticeable integration of some aspect of mental health services into the routine primary healthcare, hence improving access to these services. An emerging body of literature has reported on how implementing mental healthcare plans can effectively lead to increased integration of mental health services into primary care in low resource settings, therefore improving access and patient outcomes.^26–28^ Highlighting the improved mental health outcomes of service users after implementing the DMHPs, a previous study corroborated this by reporting that patients with depression experience a faster decrease in suicidal thoughts when treated by primary care workers (PCWs) who were part of the PRIME district mental healthcare plan in Nepal.^29^

### Implications for future research, policy and clinical practice

The findings from the study have several implications for future mental health research in Ghana. Firstly, while the current study highlights the positive impact of the DMHPs in the implementation districts, it is important to conduct further research to ascertain the long-term impact and sustainability of the plans. Longitudinal studies should be conducted to evaluate the factors that contribute to the success or failure of these plans in the long term. This can provide valuable learnings and recommendations to policy makers and healthcare workers.

One of the main challenges the study highlighted was financial constraints impacting the implementation of the DMHPs. Future research should include substantial new financial investments and assess the cost-effectiveness of the DMHPs and its various components, such as capacity building initiatives and community-based interventions, to inform resource allocation decisions. Given the limited financial resources available for mental healthcare delivery in Ghana, it is important to make informed decisions about resource allocation. Evaluating the cost-effectiveness of the different components of the DMHPs provides policy makers and implementers with important data regarding the financial feasibility of the plans for future scale-up.^30^

One of the major barriers to accessing mental healthcare in Ghana is the inadequate supply if psychotropic medicines. Revising the NHIS to include coverage for mental health services, as stipulated in the Mental Health Act 846 of 2012, would significantly improve access to care for individuals with mental health conditions. Ghana Somubi Dwumadie as part of its efforts to improve access to mental health in Ghana has been advocating for the inclusion of essential psychotropic medicines into the NHIS. It is the hope of the programme that these essential medicines would be included in the NHIS as soon as possible to improve care delivery in Ghana.

Finally, findings of the study have shown the positive impact of capacity building on mental healthcare delivery with healthcare workers reporting improved skills and knowledge in mental health detection and management. Sustaining and scaling up capacity building initiatives, such as targeted training for healthcare workers and community volunteers using the WHO mhGAP would improve the quality of mental healthcare delivery in Ghana. By investing in ongoing training, supervision, and support for healthcare workers, policymakers and healthcare facilities can ensure the long-term success and impact of DMHPs, ultimately leading to improved mental health outcomes and reducing the treatment gap in the country.

### Limitations and strengths

As limitations, the study reported from a relatively small sample size from only 3 districts from 261 districts in Ghana. While this sample size provided valuable insights, it may not cover the wide and diverse experiences of healthcare workers and service users involved in the DMHPs. The participants in this study were recruited from specific health facilities within the three demonstration districts using purposive sampling. This sampling approach, while useful for targeting relevant participants, may have introduced selection bias. The experiences and perceptions of the selected participants may not fully represent those of service users and healthcare workers in other health facilities or districts not included in the study.

The researchers involved in this study were also part of the team that implemented the DMHPs in the three demonstration districts. This dual role of the researchers as both implementers and evaluators of the program could have introduced potential biases in the data collection and interpretation processes. The researchers’ close involvement in the implementation may have influenced their perceptions and interpretations of the participants’ experiences and perspectives. Nevertheless, to mitigate this bias, the study employed reflexivity where researchers examined their biases and assumptions, as stated in the Methods section, and research assistants were employed for some data collections were not involved in the DMHP design or implementation. We observed no significant difference in the themes that emerged from research assistants’ interviews, compared to those delivered by members of the Ghana Somubi Dwumadie team (LS and KA).

Despite these limitations, the study also has some strengths. First, to our knowledge this study is the first in Ghana to explore in-depth the experiences of healthcare workers and service users in implementation of DMHPs. Using a qualitative design enabled the study team to provide a more nuanced understanding of factors supporting implementation of the DMHPs, which would have been more difficult to do using quantitative methods alone. Findings from this study have the potential to provide insights to policy makers and implementers in the scaling up of DMHPs to other districts in Ghana.

## Conclusion

The study sought the perceptions and experiences of healthcare workers and service users regarding the implementation of the DMHPs in three demonstration districts in Ghana. The findings revealed positive impacts, such as improved mental healthcare delivery through capacity building, effective collaboration, and increased awareness. Integration of mental health into primary healthcare, early identification and referral of mental health cases, and improved access to mental health services were identified as key outcomes. Service users reported reduced relapse rates, improved independence, better sleep patterns, and decreased stigma and discrimination.

However, significant challenges were also identified, including financial constraints, inadequate human resources, and shortages of psychotropic medications. Future research should assess the long-term impact, sustainability, and cost-effectiveness of the DMHPs, following more substantial financial investments. Policymakers should prioritise the inclusion of mental health services in the National Health Insurance Scheme and allocate sufficient resources to support the scale-up of DMHPs across the country. By addressing the identified challenges and leveraging the positive outcomes, Ghana can scale up the mental healthcare plan nationally to improve access to quality mental healthcare and reduce the treatment gap for individuals with mental health conditions.

## Supporting information

In-depth interview guide for healthcare workers

in-depth interview guide for service users

## Data Availability

Data are available upon reasonable request. All data relevant to the study are included in the article. Further data in the form of deidentified participant data such as interview transcripts are stored in a secure server and are available on reasonable request by emailing the corresponding author.

## Acknowledgements

This study is the result of the Ghana Somubi Dwumadie (Ghana Participation Programme). The authors express their sincere appreciation to all the participants who played a crucial role by providing data for this study.

## Contributors

Conceptualisation of study: LS, KA, LAK, BW, CL. Data acquisition: LS, KA. Data analysis: LS. Data interpretation: LS, KA, LAK, BW, CL. Drafting: LS. Critical revision: LS, KA, LAK, BW, CL. All authors have thoroughly reviewed and approved the final manuscript.

## Funding

The study was funded by the United Kingdom Foreign Commonwealth and Development Office(https://www.gov.uk/government/organisations/foreign-commonwealth-development-office) (award number: PO8604). However, the funders did not play any role in the study design, data acquisition and analysis, decision to publish, or preparation of this manuscript.

## Competing Interests

None declared.

## Ethics approval

Ethical approval was granted by the following ethics committees: King’s College London Research Ethics Committee (approval number; HR/DP-21/22-26013), and the Ghana Health Service Ethics Review Committee (approval number; GHS-ERC:006/08/21).

## Notes

### Competing Interest Statement

The authors have declared no competing interest.

### Author Declarations

Ethics committees/IRB of Kings College London and the Ghana Health Service gave ethical approval for this work

## REFERENCES

1. Arias D, Saxena S, Ephane Verguet S. Quantifying the global burden of mental disorders and their economic value. 2022; Available from: 10.1016/j.

2. Santomauro DF, Mantilla Herrera AM, Shadid J, Zheng P, Ashbaugh C, Pigott DM, et al. Articles Global prevalence and burden of depressive and anxiety disorders in 204 countries and territories in 2020 due to the COVID-19 pandemic. The Lancet [Internet]. 2021 [cited 2024 May 10]; 398:1700–12. Available from: https://www.covidminds.org/

3. Baron EC, Rathod SD, Hanlon C, Prince M, Fedaku A, Kigozi F, et al. Impact of district mental health care plans on symptom severity and functioning of patients with priority mental health conditions: The Programme for Improving Mental Health Care (PRIME) cohort protocol. BMC Psychiatry [Internet]. 2018 Mar 6 [cited 2024 Jun 4];18(1):1–14. Available from: https://bmcpsychiatry.biomedcentral.com/articles/10.1186/s12888-018-1642-x

4. Ae-Ngibise KA, Sakyi L, Adwan-Kamara L, Lund C, Weobong B. Prevalence of probable mental, neurological and substance use conditions and case detection at primary healthcare facilities across three districts in Ghana: findings from a cross-sectional health facility survey. BMC Psychiatry [Internet]. 2023 Dec 1 [cited 2024 May 30];23(1):1–12. Available from: https://bmcpsychiatry.biomedcentral.com/articles/10.1186/s12888-023-04775-z

5. Weobong B, Ae-Ngibise KA, Sakyi L, Lund C. Towards implementation of context-specific integrated district mental healthcare plans: A situation analysis of mental health services in five districts in Ghana. PLoS One. 2023 May 1;18(5 May).

6. Patel V, Saxena S, Lund C, Thornicroft G, Baingana F, Bolton P, et al. The Lancet Commission on global mental health and sustainable development. Vol. 392, The Lancet. Lancet Publishing Group; 2018. p. 1553–98.

7. Lund C, Tomlinson M, Patel V. Integration of mental health into primary care in low- and middle-income countries: the PRIME mental healthcare plans. The British Journal of Psychiatry [Internet]. 2016 Jan 1 [cited 2024 May 11];208(Suppl 56):s1. Available from: /pmc/articles/PMC4698550/

8. Hoeft TJ, Fortney JC, Patel V, Unützer J. Task Sharing Approaches to Improve Mental Health Care in Rural and Other Low Resource Settings: A Systematic Review. J Rural Health [Internet]. 2018 Dec 1 [cited 2024 May 11];34(1):48. Available from: /pmc/articles/PMC5509535/

9. Luitel NP, Jordans MJD, Subba P, Komproe IH. Perception of service users and their caregivers on primary care-based mental health services: A qualitative study in Nepal. BMC Fam Pract [Internet]. 2020 Sep 28 [cited 2024 Jun 26];21(1):1–11. Available from: https://bmcprimcare.biomedcentral.com/articles/10.1186/s12875-020-01266-y

10. Staab EM, Wan W, Li M, Quinn MT, Campbell A, Gedeon S, et al. Integration of Primary Care and Behavioral Health Services in Midwestern Community Health Centers: A Mixed Methods Study. Families, Systems and Health. 2022;40(2):182– 209.

11. WHO. mhGAP intervention guide for mental, neurological and substance use disorders in non-Specialized health settingslJ: mental health gap action programme (mhGAP). 2016.

12. Ae-Ngibise KA, Sakyi L, Adwan-Kamara L, Weobong B, Lund C. Development and implementation of mental healthcare plans in three districts in Ghana: a mixed method process evaluation using the MRC Complex Interventions framework and Theory of Change. Preprint [Internet]. 2024; Available from: 10.21203/rs.3.rs-3826045/v1

13. Madlala ST, Miya RM, Zuma M. Experiences of mental healthcare providers regarding integration of mental healthcare into primary healthcare at the Ilembe Health District in Kwazulu-Natal Province. Health SA Gesondheid. 2020;25:1–8.

14. Braun V, Clarke V. Using thematic analysis in psychology. Qual Res Psychol. 2006;3(2):77–101.

15. Azungah Theophilus. Qualitative research: deductive and inductive approaches to data analysis. Qualitative Research Journal. 2018;18(4):383–400.

16. Pereira B, Andrew G, Pednekar S, Kirkwood BR, Patel V. The integration of the treatment for common mental disorders in primary care: Experiences of health care providers in the MANAS trial in Goa, India. Int J Ment Health Syst. 2011 Oct 3;5.

17. Kiima D, Jenkins R. Open Access SHORT REPORT Mental health policy in Kenya-an integrated approach to scaling up equitable care for poor populations [Internet]. Vol. 4, International Journal of Mental Health Systems. 2010. Available from: http://www.ijmhs.com/content/4/

18. Hijazi Z, Weissbecker I, Chammay R. The integration of mental health into primary health care in Lebanon. Intervention (Amstelveen). 2011;9:265–78.

19. Petersen I, Marais D, Abdulmalik J, Ahuja S, Alem A, Chisholm D, et al. Strengthening mental health system governance in six low- and middle-income countries in Africa and South Asia: Challenges, needs and potential strategies. Health Policy Plan. 2017 Jun 1;32(5):699–709.

20. Luitel NP, Jordans MJD, Adhikari A, Upadhaya N, Hanlon C, Lund C, et al. Mental health care in Nepal: Current situation and challenges for development of a district mental health care plan. Confl Health. 2015;9(1).

21. Hanlon C, Luitel NP, Kathree T, Murhar V, Shrivasta S, Medhin G, et al. Challenges and opportunities for implementing integrated mental health care: a district level situation analysis from five low- and middle-income countries. PLoS One [Internet]. 2014 Feb 18 [cited 2024 Jun 18];9(2). Available from: https://pubmed.ncbi.nlm.nih.gov/24558389/

22. Njau T, Mwakawanga DL, Sunguya B, Minja A, Kaaya S, Fekadu A. Perceived barriers and opportunities for implementing an integrated psychological intervention for depression in adolescents living with HIV in Tanzania. BMC Health Serv Res [Internet]. 2024 May 28 [cited 2024 Jun 18];24(1):672. Available from: http://www.ncbi.nlm.nih.gov/pubmed/38807134

23. Bhana A, Petersen I, Baillie KL, Flisher AJ. Implementing the World Health Report 2001 recommendations for integrating mental health into primary health care: a situation analysis of three African countries: Ghana, South Africa and Uganda. Int Rev Psychiatry [Internet]. 2010 Dec [cited 2024 Jun 18];22(6):599–610. Available from: https://pubmed.ncbi.nlm.nih.gov/21226648/

24. Ghana Somubi Dwumadie. Policy brief Access to psychotropic medicines in Ghana: Issues, strategies, and recommendations Key messages [Internet]. 2023. Available from: https://www.who.int/publications/m/item/mental-health-atlas-gha-2020-country-profile

25. Shidhaye R, Baron E, Murhar V, Rathod S, Khan A, Singh A, et al. Community, facility and individual level impact of integrating mental health screening and treatment into the primary healthcare system in Sehore district, Madhya Pradesh, India Handling editor Seye Abimbola. BMJ Glob Health. 2019;4:1344.

26. Petersen I, Ssebunnya J, Bhana A, Baillie K. Lessons from case studies of integrating mental health into primary health care in South Africa and Uganda. Int J Ment Health Syst. 2011 Apr 15;5.

27. Shidhaye R, Shrivastava S, Murhar V, Samudre S, Ahuja S, Ramaswamy R, et al. Development and piloting of a plan for integrating mental health in primary care in Sehore district, Madhya Pradesh, India. British Journal of Psychiatry. 2016 Jan 1;208:s13–20.

28. Wolff LS, Flynn A, Xuan Z, Errichetti KS, Tapia Walker S, Brodesky MK. The effect of integrating primary care and mental health services on diabetes and depression: A multi-site impact evaluation on the US-Mexico Border. Med Care. 2021 Jan 1;59(1):67–76.

29. Aldridge LR, Garman EC, Luitel NP, Jordans MJD. Impact of a district mental health care plan on suicidality among patients with depression and alcohol use disorder in Nepal. PLoS One. 2020;15(4).

30. Chisholm D, Burman-Roy S, Fekadu A, Kathree T, Kizza D, Luitel NP, et al. Estimating the cost of implementing district mental healthcare plans in five low-and middle-income countries: The PRIME study. British Journal of Psychiatry. 2016 Jan 1;208:s71–8.

