## Supplementary material for "Care delivery in the context of district mental healthcare plans (DMHP) in Ghana: experiences of primary health care workers and service users": In-depth interview guide for healthcare workers

**In-depth Interviews**

A. **Health care workers and Mental Health Care Plan (MHCP) implementers**

**Introduction**

Thank you for agreeing to participate in this research interview. The purpose of this discussion is to explore your views about the acceptability, feasibility, and process of implementing the MHCPs in your district. We want to understand what works with regards to implementing the MHCP in your district. In your experience as a (service provider, other key stakeholders), we would like to explore with you: what worked well, the challenges you encountered and what needs to be done differently to improve delivery of mental health services in your district.

a. **Lessons learned from implementing the MHCP:**
1. What key lessons were learnt from your experience in implementing the MHCP in your district? Probe for overall improved service delivery and supervision, feasibility, and acceptability etc.
b. **Acceptability of delivering mental healthcare in primary care:**
1. In your assessment of the MHCP implementation so far, do you think this plan is acceptable by various stakeholders including the following?
   a. Healthcare providers? explain
   b. Services users? explain
   c. Community members? explain?
c. **Availability resources for implementation of the MHCP:**
1. What are the resources needed for implementing the MHCP in your district? Probe further for:
a. Physical resources (egg physical structures and space, medication, access and coverage of mental health services, means of transport for community outreaches etc)
b. Human resources (erg adequate number of health personnel for delivering services, training and supervision etc)
c. Financial resources (erg mental health budget in place)
2. What is your assessment of implementing the MHCP in your district? What works very well in your estimation? Probe for:
a. increased number of people correctly receiving evidence-based treatment?
b. Improved health, social and economic outcomes of people living with priority mental disorders?
c. Increased coverage of evidence-based mental health services?
3. What did not work so well? Explain and give reasons
4. What can be done better or differently (or what strategies will you propose) to expedite implementation of the MHCP in your district? Please discuss and provide examples.
d. **Funding for district MHCP implementation:**
1. How well are the different parts of the MHCP implemented in your district?
2. What is the estimated cost for implementing such a plan in the district?
e. **Political and socio-cultural determinants:**
1. How committed is the state in ensuring resources are made available for implementation of the MHCP in your district? Please discuss. (Policies & programmes, commitment of resources for mental health care, training, etc.)
2. To what degree do cultural belief systems and mental health literacy play a role in implementation of the MHCP? Please discuss
3. What are the cultural determinants of implementing the MHCP in your district?
f. **Social Protection as economic empowerment**

From your experience:

1. Through the implementation of the MHCP, are persons with mental health disorders registered with the Social Welfare Department for the Livelihood Empowerment Against Poverty entitlement? Please discuss – what were the challenges you encountered in getting people registered? How were those challenges overcome?
2. Are people with mental health conditions registered for National Health Insurance Scheme? Please discuss. – what were the challenges you encountered in getting people registered? How were those challenges overcome?
g. **Training and Supervision:**
1. Do you feel all the implementers of the MHCP are adequately trained for the implementation of the plan? discuss
2. From your experience, how effective is supervision for implementing the MHCP in your district?

**General comments:**

1. What is the level of progress achieved with overall implementation of the MHCP? Please discuss.
2. What do you see as the major challenges in implementation of the MHCP in your district? Please discuss.
3. Do you have anything else to add to our discussion today regarding how well the MHCP has been implemented?
4. Sustainability: do you think the MHCP will continue to be implemented? What will help to make this happen? What barriers will you be likely to encounter and how might these be overcome?
