## Supplementary material for "Care delivery in the context of district mental healthcare plans (DMHP) in Ghana: experiences of primary health care workers and service users": in-depth interview guide for service users

A. **In-depth Interviews – Mental health services users**
B. **Introduction**

Thank you for agreeing to participate in this research interview. District Mental Health Care Plans are comprehensive plans that aim to address the mental health needs of individuals in this district. The main goal of District Mental Health Care Plans is to improve access to quality mental health care services and support for individuals experiencing mental health challenges. As part of the strategy to improve access to mental healthcare services in Ghana, the programme is facilitating the implementation of District Mental Health Care Plans (DMHCPs). The purpose of this discussion is to explore your views about the acceptability of implementing the MHCPs in your district, barriers adhering to protocol treatment as well as experience of stigma and discrimination. In your experience as a service user, we want to understand what works with regards to implementing the MHCP in your district.

C. **Mental health services users**
a. **Acceptability of the health care provided:**
1. In your experience, will you say that the services you received from health care providers are acceptable?
2. How satisfied were you with the services you received during your last visit to the health care provider? Probe for quality, timeliness, and effectiveness etc
3. In your experience, how acceptable and feasible is the MHCP being implemented in your district?
4. Has the introduction of the MHCP resulted in access to affordable mental healthcare services? Please discuss.
5. Has the introduction of the MHCP increased availability of community-based mental health services (that is mental health services that are not provided in hospitals but in the community, close to the places most people live and work)? Please discuss and provide examples.
b. **Barriers adhering to treatment protocol:**
1. What do you think are the barriers in adhering to treatment? Probe for lack of medication, distance to service provision, affordable services, etic
2. What can be done to improve adherence to treatment?
c. **Experience of stigma and discrimination:**
1. Has the introduction and implementation of the MHCP contributed to a reduction in stigma and discrimination against people with mental disorders? Please discuss and provide examples. Did it unintentionally lead to an increase in stigma and discrimination? If so, how?
2. Do you feel more inclusive in your community and family life following the introduction and implementation of the MHCP in your district? Explain
d. **Relationships between treatment and functional and economic status:**
1. Have your health condition improved sense the implementation of the MHCP in your district?
2. Are you able to do the day-to-day activities that are important to you?
3. Will you say you are economically productive now than before you received treatment? Will you attribute your current economic status to the treatment you received following the introduction of the MHCP?
e. **Collaboration between traditional and faith-based healers:**
1. What is the role of traditional and faith-based healers in delivering mental health services in your district?
2. What is your experience regarding the quality of collaboration between health care providers and traditional and faith-based healers?
3. What can be done to improve this collaboration and improve case management in your district?
